# Depressive symptoms and single nucleotide polymorphisms predict clinical recurrence of inflammatory bowel disease

**DOI:** 10.1101/2020.06.29.20139030

**Authors:** Sebastian Bruno Ulrich Jordi, Brian Matthew Lang, Bianca Auschra, Roland von Känel, Luc Biedermann, Thomas Greuter, Philipp Schreiner, Gerhard Rogler, Niklas Krupka, Michael Christian Sulz, Benjamin Misselwitz, Stefan Begré, on behalf of the Swiss IBD cohort study group

## Abstract

**Background and Aims:** Inflammatory bowel disease (IBD) patients are at high risk for depression. We examined interrelations between genetic risk factors for depression, depressive symptoms and IBD flares.

**Methods:** In 1973 patients (1137 Crohn’s disease, 836 ulcerative colitis) of the Swiss IBD cohort study (SIBDC), 62 single nucleotide polymorphisms (SNPs) preselected for associations with depression, stress, pain and smoking were screened for cross-sectional associations with depression (hospital anxiety and depression subscale for depression, HADS-D≥11). Logistic regression and Cox proportional hazards models were built to test for effects of depressive symptoms on disease course and genetic risk factors on depression and disease course. As endpoints we used active disease (CDAI≥150 or MTWAI≥10) and two published composite flare definitions: FNCE: physician reported flare, non-response to therapy, new complication or extraintestinal manifestation and AFFSST: active disease, physician reported flare, fistula, stenosis and new systemic therapy.

**Results:** Depressive symptoms were a strong risk factor for disease related endpoints including active disease (adjusted hazard ratio, aHR: 3.25, p<0.001), AFFSST (aHR: 1.62, p<0.001) and FNCE (aHR: 1.35, p=0.019). Rs588765’s TC alleles and rs2522833’s C allele were associated with depressive symptoms at baseline (odds ratio, OR: 0.43, q=0.050 and OR: 1.73, q=0.059, respectively). Rs588765-TC remained protective regarding presence of depression (aHR: 0.67, p=0.035) and was associated with fewer active disease states (aHR: 0.72, p=0.045) during follow-up.

**Conclusions:** In IBD, genetics, depressive symptoms and inflammatory activity are intimately related: Depressive symptoms were a predictor of clinical deterioration and rs588765-TC was protective for depression and high IBD activity.

**Funding:** This work was supported by the Swiss National Science Foundation (SNSF).

## Introduction

Inflammatory bowel diseases (IBD) are frequent conditions affecting approximately 2.2 million people in Europe and 1.5 million Americans with an increasing incidence worldwide.^1^ IBD, comprising the subtypes Crohn’s disease (CD), ulcerative colitis (UC) and indeterminate colitis (IC) are immunologically mediated diseases which can severely impair affected individuals’ lives. The disease course of IBD is variable with periods of quiescent disease which are interrupted by IBD flares, i.e. episodes of more active inflammation.

The exact cause of IBD remains elusive, but likely involves an interplay of genetic, microbial, immunological and environmental factors.^2^ A significant part of IBD risk is heritable and more than 230 single nucleotide polymorphisms (SNP) associated with IBD have been identified.^3^ These SNPs partially explain an individua’s vulnerability to develop IBD but have not been useful to predict the clinical disease course in IBD patients.^4^ Many environmental factors have been implicated as trigger factors for IBD flares including diet, medication and smoking.^5^

Psychiatric conditions such as stress, depression and anxiety have been associated with IBD and the risk to experience depression is higher for IBD patients compared to healthy individuals (incidence rate ratio: 1.6).^6^ Furthermore, stress and depression overlap with pain^7^, pain sensitivity^8^ and smoking.^9^ Stress, depression and anxiety have been suggested as triggers of flares and clinical deterioration in IBD.^10,11^

Depressive disorders are heritable conditions with heritability rates of approximately 40%^12^ and many individual SNPs have been associated with the risk for depression.^13^ An example for a possible genetic origin of the observed overlap of depression, smoking and pain sensitivity are neuronal nicotinic acetylcholine receptors (nAChRs). Recent findings suggest that nAChRs are involved in psychiatric disorders such as depression although mechanistic details remain unclear.^14^ Moreover, nAChRs had been associated with nicotine and alcohol dependence,^15,16^ also among SIBDC patients.^17^ Currently, nAChRs are investigated as pharmacological targets for depression, anxiety, pain and nicotine addiction.^18^ For these reasons, SNPs within nAChR genes might influence the occurrence of flares in IBD via their association with depression and/ or their link to smoking.

However, the role of genetic risk factors for depressive symptoms has not yet been clarified for individuals at high risk for depression such as IBD patients and also the interplay between genetic and environmental risk factors of depression remains unclear.

We are using data from the Swiss IBD cohort study (SIBDC)^19^ to explore the relationship between depression, individual SNPs and the risk for IBD deterioration. SIBDC was started in 2006 as a prospective cohort study that recruits nationwide in Switzerland.^19,20^ The impact of depressive symptoms, anxiety and social support on IBD flares has been studied in SIBDC before;^10,21^ we were aiming to validate, generalize and extend these findings, and add a genetic context.

## Methods

### Design of the SIBDC

The SIBDC was started in 2006 as a prospective cohort study that recruits nationwide in Switzerland.^19,20^ New patients are continuously recruited and patients are followed yearly. IBD-related clinical data is collected by physicians at enrolment and yearly follow-ups. In addition, clinical, sociodemographic and psychosocial data are acquired using patient questionnaires at enrolment and yearly follow-ups. All SIBDC patients consenting to genetic analysis with appropriate biomaterial available were genotyped and only genotyped patients were included in our study.

SIBDCS is funded by the Swiss National Science Foundation and has been approved by the local ethics committee of each participating center (institutional review board approval No. EK-1316, approved on February 5, 2007 and KEK Zurich, March 9, 2020; BASEC 2018-02068). SIBDC goals and methodology are described elsewhere.^19,20^ All patients provided written informed consent prior to inclusion into the SIBDCS. Analysis of patient data for the current study was approved by the scientific board of SIBDCS.

### Patient characteristics

We exported clinical and sociodemographic variables acquired at enrolment and follow-ups: Disease activity was measured using the Crohn’s disease activity index CDAI or the modified Truelove and Witts activity index (MTWAI) for CD and UC/IC patients, respectively. For intercomparable measures, CDAI or MTWAI score values were Z-score normalized to a disease activity score 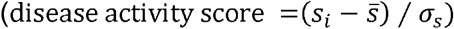 or cutoff values for active disease were used (see below). The following binary variables were created by distinguishing between patients that had any of the respective included criteria and patients that did not: *Complications* included colorectal cancer, colon dysplasia, intestinal lymphoma, osteopenia/osteoporosis, anemia (not as an adverse event of medical therapies), deep vein thrombosis, pulmonary embolism, nephrolithiasis, gallstones,^22^ malabsorption syndrome, massive hemorrhage, perforation or peritonitis. *Extraintestinal manifestations* (EIM) comprised peripheral arthritis/arthralgia, uveitis/iritis, pyoderma gangrenosum, erythema nodosum, aphthous oral ulcers/stomatitis, ankylosing spondylitis/sacroiliitis and primary sclerosing cholangitis (PSC). Any stenosis localized in the esophagus, duodenum/jejunum, ileum, large bowel, rectum or anus was summarized in the *stenosis* variable. The *fistula* variable comprised IBD-related fistula and abscesses and anal fissure, whereas the variable *surgery* included any abdominal or fistula and abscess-related surgery. *TNF inhibitors* referred to treatments with infliximab, certolizumab pegol or adalimumab (no patient with golimumab treatment was reported at the time of data export). The variable *number of current therapies* was created by counting presently administered therapies which could be any of the following: any steroids, any antibiotics, 5-aminosalicylic acid, sulfasalazine, azathioprine, 6-mercaptopurine, methotrexate, cyclosporine, tacrolimus, infliximab, adalimumab, certolizumab pegol, cholestyramine, *E. coli* Nissle (Mutaflor^®^), ursodeoxycholic acid, bisphosphonates, the probiotic VSL#3^®^, or any other medication that was noted by physicians. A patient was considered a smoker when he or she declared to have a current habit of smoking independent of the quantity.

### Outcome measures

Depressive symptoms were measured with the Hospital Anxiety and Depression Scale’s (HADS) subscale for depression (HADS-D).^23^ Following well-established criteria, the presence of depression was defined as HADS-D ≥11, indicating probable moderate or severe depression.^24-26^ This was done for both a constant (at baseline) and a time-varying depression variable. In this study, the term *depression* is used with reference to this definition unless stated otherwise.

Active disease was defined by CDAI ≥150^27^ or MTWAI ≥10^28^, respectively. An IBD flare was measured as a composite endpoint of clinical variables. IBD is a complex disease and we considered two different flare definitions, following the examples of two prior SIBDC studies.^10,21^ The first composite clinical endpoint ***a****ctive disease, physician reported* ***f****lare*, ***f****istula, stenosis*, ***s****urgery or new* ***s****ystemic* ***t****herapy* (AFFSST) was reached upon occurrence/persistence of active disease, defined by CDAI ≥150^27^ or MTWAI ≥10^28^, a physician reported flare, new fistula, new stenosis, new surgery, intake of systemic steroids and/or start of therapy with a new TNF inhibitor (also changes between TNF inhibitors) at follow-up.^10^

The second composite clinical endpoint *physician reported* ***f****lare*, ***n****on-response to any administered therapy with consequent transition to a more aggressive medication, new complication or new* ***E****IM* (FNCE) was defined as the occurrence of any one of these events at a follow-up examination.^21^

For both composite endpoints we counted the first occurrence of each defining feature after enrolment as event. Both composite endpoints were reached upon the first occurrence of any of these events.

For data analyses with survival techniques, all endpoints (the individual flare features, the FNCE and AFFSST composite endpoints and depression) were coded with right censoring. Thereby, a time interval t_n-1_-t_n_ was defined where t_n_ corresponds to the n^th^ follow-up visit when the event was documented first. This way, we describe a time interval during which an event had occurred [t_n-1_, t_n_] and then assumed an event’s occurrence at t_n-1_+(t_n_-t_n-1_)/2 on average.

### Selection of candidate SNPs

Allelic state of a number of SNPs were determined in SIBDC patients. We selected all 95 SNPs available without a link to inflammation or IBD diagnosis. These *non-inflammatory SNPs* had been selected with three objectives: 48 SNPs were selected for an association with stress and/or depressive symptoms, based on a thorough literature search in PubMed and SNPedia. Moreover, 30 SNPs with a suspected association with pain were included. Furthermore, 16 SNPs associated with smoking in all 3 genome wide association studies available in 2015 were included.^17,29-31^ Both, pain^7^ and smoking^11,37^ have a strong reciprocal associations with depression and anxiety.

For calculation of linkage disequilibrium (LD) we used the square of the correlation coefficient (r^2^) between two SNPs. Groups were formed whenever two or more SNPs shared an r^2^ value of 0.5 or higher, representing moderate LD.^32^ The SNP with the strongest correlation to group members was chosen as the representing tag-SNP for the respective group. If there were only two SNPs in a group, the SNP with the lowest number of missing data points in the respective group was selected as tag SNP.

### Screening for depression-associated SNPs

For the screening of SNPs associated with depression, we used methods of the *SNPassoc* R package^33^ version 1.9-2, developed and established for whole genome association studies. We calculated the association between each of our SNP and depression while simultaneously selecting the genetic model (codominant, dominant, recessive, overdominant, log-additive) for each SNP which best describes this relationship via likelihood ratio test comparison. We filtered the list of SNPs to those with a Bonferroni corrected p-value (q-value) <0.1. Associations were confirmed by multivariable analyses with depressive symptoms at enrolment as endpoint. For multivariable analyses with depressive symptoms as dependent variable in this study, a set of control variables was defined (set 1: *sex, diagnosis, age, time since diagnosis, BMI, smoking status, alcohol consumption, standardized CDAI/MTWAI score, abdominal surgery prior to enrolment, systemic steroids, TNF inhibitors, number of current therapies*). These variables were tested for association with depressive symptoms at enrolment using univariable logistic regression analysis.

Models included all control variables and were calculated for every SNP individually. To minimize the risk of obtaining significance due to overfitting we also created a simpler secondary model by automated variable elimination. In this process the optimal model regarding the AIC was calculated. For this we used the *glmulti* R package^34^ version 1.0.7.1.

### Time-to-event analyses

To confirm associations with SNPs at enrolment (cross-sectional data), we constructed time-to-event models using the *Survival* R package^35^ version 3.1-12. We implemented univariable and multivariable Cox proportional hazards models with the endpoint of interest being presence of depressive symptoms (HADS ≥11) at a follow-up examination, independent of a patients HADS score value at enrolment.

Clinical disease course was described by the absence or presence of either active disease or flares according to composite endpoint definitions (see above). To assess the impact of depressive symptoms and depression-associated SNPs on the clinical course of IBD,^10^ we coded a time-varying control variable for depression (HADS-D score ≥11) that reflected a patient’s depression status over time.^36^ To ensure an unambiguous time sequence of time-varying depression status and clinical endpoint measurements (e.g. flares), a conservative coding approach was chosen: only if a recording of depressive symptoms preceded a flare, an association was made. Depression status was defined forwardly, while flares were defined backwardly. For example, measures for depression at time point t_n_ defined the time-varying depression variable for the interval t_n_ until t_n+1_, while recordings of clinical variables at time point t_n_ defining a clinical endpoint variable (e.g. flare) where assumed to have happened at the time point in the middle between the last and the current follow-up (calculated as t_n-1_+(t_n_-t_n-1_)/2). As a result, concurrent depression and flares will not be detected as an association.

For these time-to-event analyses with clinical endpoints we adapted the set of control variables by removing variables that could be proxy-variables for clinical endpoints (set 2: *sex, diagnosis, time since diagnosis, age, BMI, disease related surgery prior to enrolment, smoking status, daily alcohol consumption*). As before, univariable and multivariable Cox proportional hazards models were implemented. All calculations were performed using *R*^37^ version 3.6.1.

## Results

### Study participants

For our analysis, data from 1973 genotyped SIBDC patients (1137 with CD, 836 with UC/IC) were available with regular follow-ups up to 4250 days. Baseline characteristics of our cohort sample are presented in **Table 1**, confirming a mixed study population with mild, moderate and severe disease. Depression (HADS-D ≥11) at enrolment tended to be more prevalent in CD compared to UC (9.5% in CD vs. 6.5% in UC/IC, p=0.052), consistent with earlier findings.^38^ Smoking,^39^ fistulae, stenosis, EIMs, complications, previous abdominal surgery and therapy with TNF inhibitors was more common in CD patients and CD patients had significantly longer disease courses and lower BMIs. On the other hand, UC/IC patients had a higher number of different therapies at enrolment and were more often treated with systemic steroids.

**Table 1:**
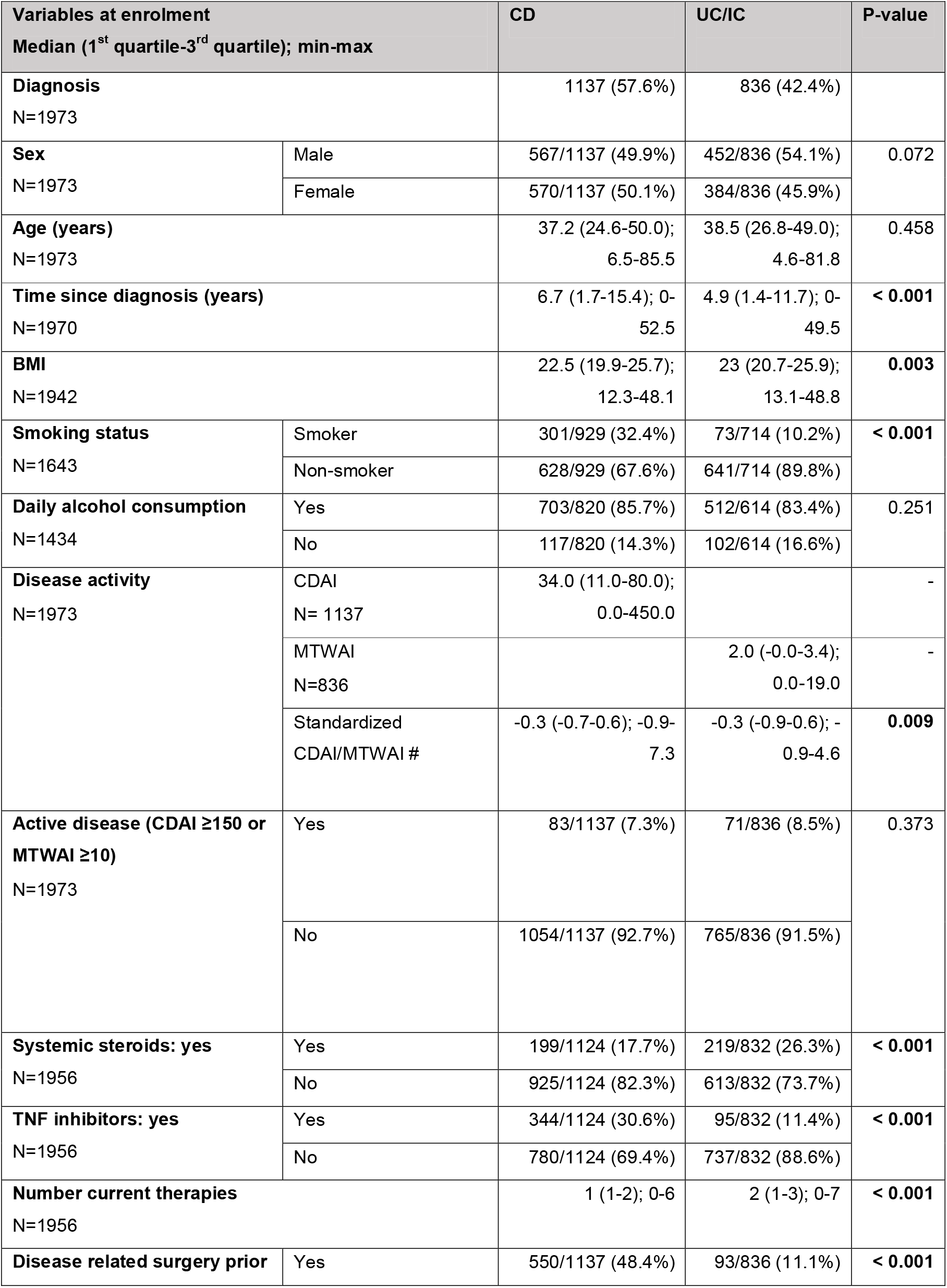
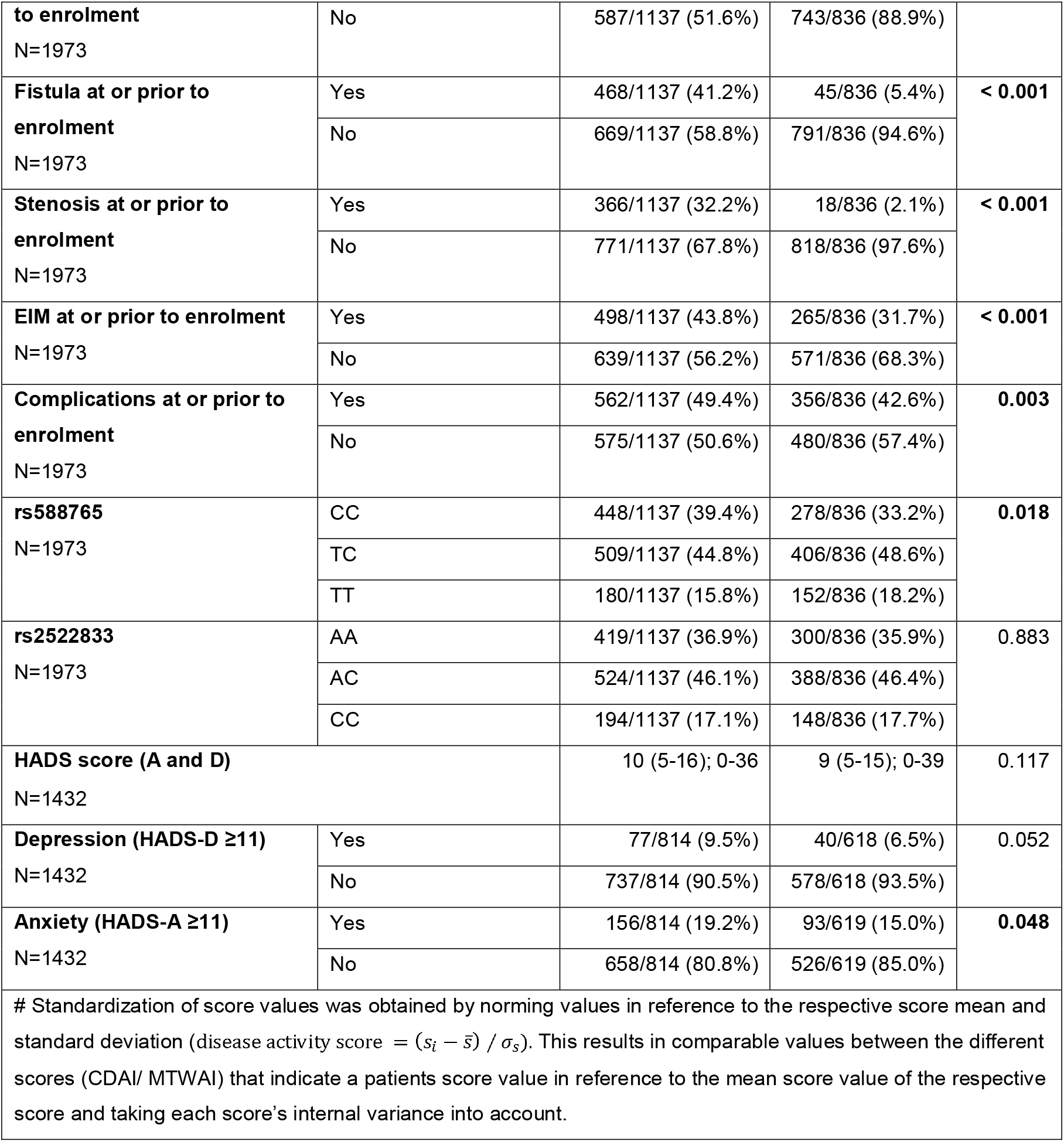
Study participants. Characteristics of study participants at enrolment stratified for diagnosis. Abbreviations: CD: Crohn’s disease, UC: ulcerative colitis, IC: indeterminate colitis, BMI: body mass index, CDAI: Crohn’s Disease Activity Index, MTWAI: Modified Truelove and Witts Severity Index, TNF: tumor necrosis factor, T: thymine, C: cytosine, A: adenine, HADS: Hospital Anxiety and Depression Scale, HADS-D: subscale for depressive symptoms, HADS-A: subscale for anxiety symptoms.

Depressive symptoms at enrolment were significantly associated with several control variables including BMI (p=0.01), daily alcohol consumption (p=0.017), disease activity (p<0.001) and number of current therapies (p=0.004, **Table 2)**.

**Table 2.** Univariable logistic regression analyses - odds ratios for depression in IBD patients. **Variables associated with depression (HADS-D ≥11) at baseline.** Univariable logistic regression models showing the association of possible confounders for multivariable models with depression. Results are presented as odds ratios; p-values are indicated as follows: *: p<0.1, **: p<0.05, ***: p<0.01. 1043 patients were analyzed. Abbreviations: CI: 95% confidence interval, IBD: inflammatory bowel disease, HADS: Hospital Anxiety and Depression Scale, UC: ulcerative colitis, IC: indeterminate colitis, CD: Crohn’s disease, BMI: body mass index, CDAI: Crohn’s Disease Activity Index, MTWAI: Modified Truelove and Witts Severity Index, TNF: tumor necrosis factor.

|  |  |
| --- | --- |
| **Sex: male** | 0.91 (CI: 0.58-1.43) |
| **Diagnosis: UC/IC (vs. CD)** | 0.79 (CI: 0.50-1.26) |
| **Age (years)** | 1.01 (CI: 1.00-1.03) |
| **Time since diagnosis (years)** | 1.00 (CI: 0.97-1.02) |
| **BMI** | **1.07 (CI: 1.02-1.11) \*\*\*** |
| **Smoking status: smoker** | 1.42 (CI: 0.90-2.26) |
| **Daily alcohol consumption: yes** | **3.12 (CI: 1.24-7.83) \*\*** |
| **Disease activity index (standardized CDAI/MTWAI)** | **1.80 (CI: 1.53-2.13) \*\*\*** |
| **Disease related surgery prior to enrolment** | 1.00 (CI: 0.64-1.62) |
| **Systemic steroids: yes** | 1.55 (CI: 0.95-2.54) * |
| **TNF inhibitors: yes** | 1.30 (CI: 0.78-1.59) |
| **Number current therapies** | **1.01 (CI: 1.00-1.03) \*\*\*** |

### Screening of SNPs for an association with depressive symptoms

SNPs were tested for linkage disequilibrium **(Supplementary figure 1)** and SNPs with a pair-wise correlation coefficient r^2^ ≥0.5 aggregated in 16 groups. From each group only one representative SNP was used henceforward, reducing the number of SNPs from 95 to 62 **(Supplementary table 1)**.

**Figure 1:**
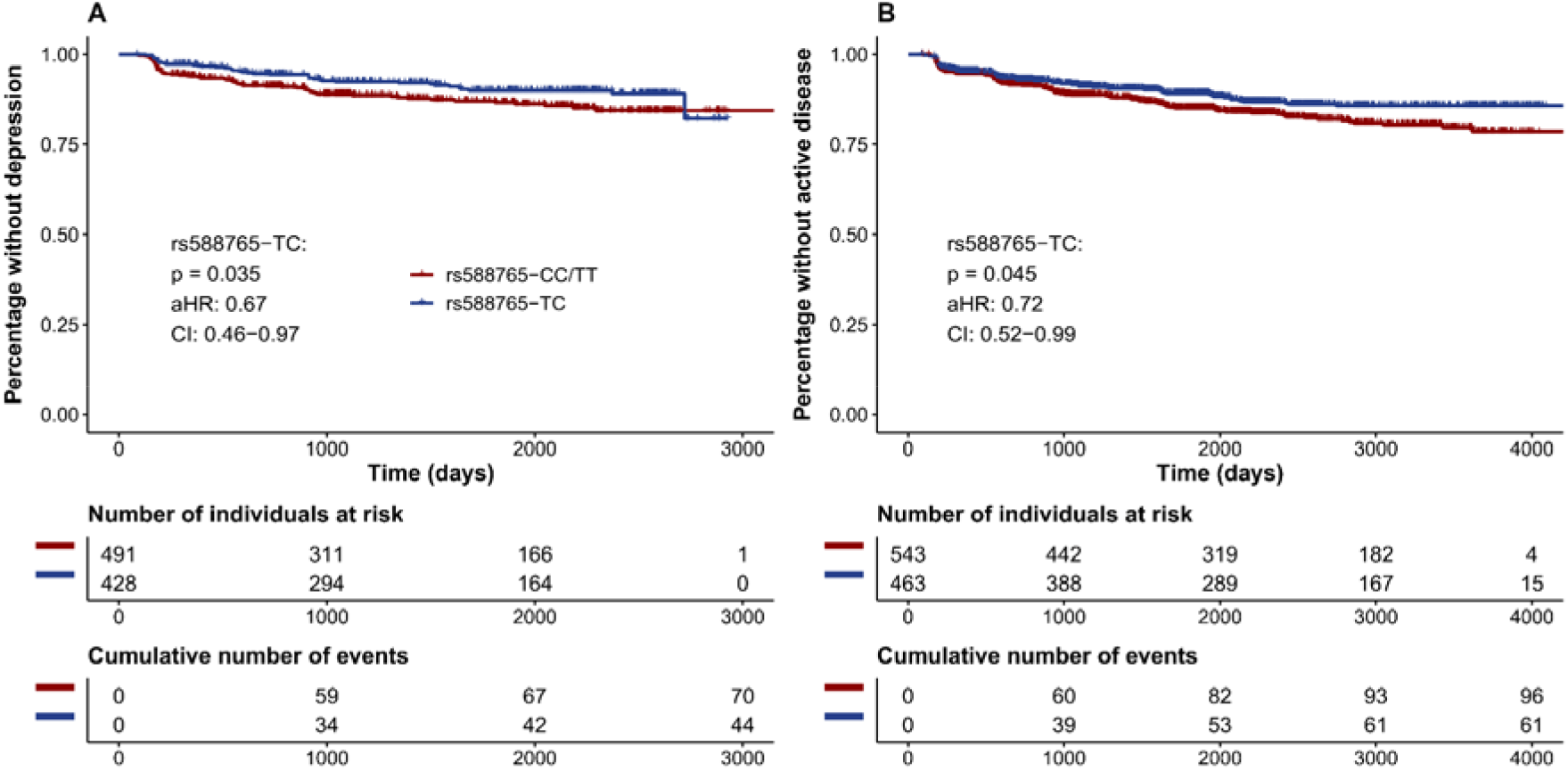
Kaplan Meier curves showing how rs588765’s genotype impacts the hazards for depression and IBD activity over time. **A:** Percentage of IBD patients without depression stratified for rs588765 genotype (overdominant). **B:** Percentage of IBD patients without active disease defined as CDAI ≥150 or MTWAI ≥10 in CD and UC patients, respectively and stratified for rs588765 genotype (overdominant). Analysis: multivariable Cox proportional hazards models. Abbreviations: HR: hazard ratio, CI: confidence interval, T: thymine, C: cytosine, IBD: inflammatory bowel disease, CDAI: Crohn’s Disease Activity Index, MTWAI: Modified Truelove and Witts Severity Index.

All SNPs were screened for an association with depression (HADS-D ≥11) at enrolment in three different models: univariable, complete multivariable and reduced multivariable model after automated parameter elimination **(Supplementary table 2)**. Of 62 SNPs tested, two SNPs, rs588765 and rs2522833, were significantly associated with depressive symptoms at enrolment with q<0.1 (Bonferroni corrected). In the univariable model, for rs2522833 the association was strongest in a log-additive inheritance model with the C allele as an associated risk allele (odds ratio, OR: 1.71, CI: 1.25-2.34, q=0.043). For rs588765, an overdominant genetic pattern yielded the strongest significance identifying TC as an associated protective allele combination (OR: 0.45, CI: 0.28-0.74, q=0.066, **Table 3** and **Supplementary figure 2**). For both SNPs, the associations detected in the univariable models stayed significant (q<0.1) in multivariable models correcting for possible confounders (rs2522833: q=0.045; rs588765: q=0.061, **Supplementary figure 3)**, also after parameter elimination (rs2522833: q=0.059; rs588765: q=0.050, **Supplementary figure 4**). This indicates a robust association between both SNPs and depressive symptoms

**Table 3.** Association analysis - odds ratios for depression in IBD patients. **SNPs associated with depression (HADS-D ≥11).** Table showing the analysis results of the two SNPs significantly (q<0.1) associated with depression at enrolment in IBD patients. Results are presented as odds ratios; q-values are indicated as follows: *: q<0.1, **: q<0.05, ***: q<0.01. Analyses: logistic regression analysis. Abbreviations: CI: 95% confidence interval, IBD: inflammatory bowel disease, HADS: Hospital Anxiety and Depression Scale, T: thymine, C: cytosine.

| Dependent variable: depression at enrolment (HADS depression sub score $\geq 11$) | | | |
| --- | --- | --- | --- |
|  | Univariable | Multivariable without variable elimination | Multivariable with variable elimination |
| rs588765: TC (vs. CC/TT) | 0.45 (CI: 0.28-0.74) * | 0.43 (CI: 0.26-0.72) * | 0.43 (CI: 0.25-0.72) ** |
| rs2522833: number of C alleles (0-2) | 1.71 (CI: 1.25-2.34) ** | 1.75 (CI: 1.26-2.44) ** | 1.73 (CI: 1.25-2.39) * |

**Figure 2:**
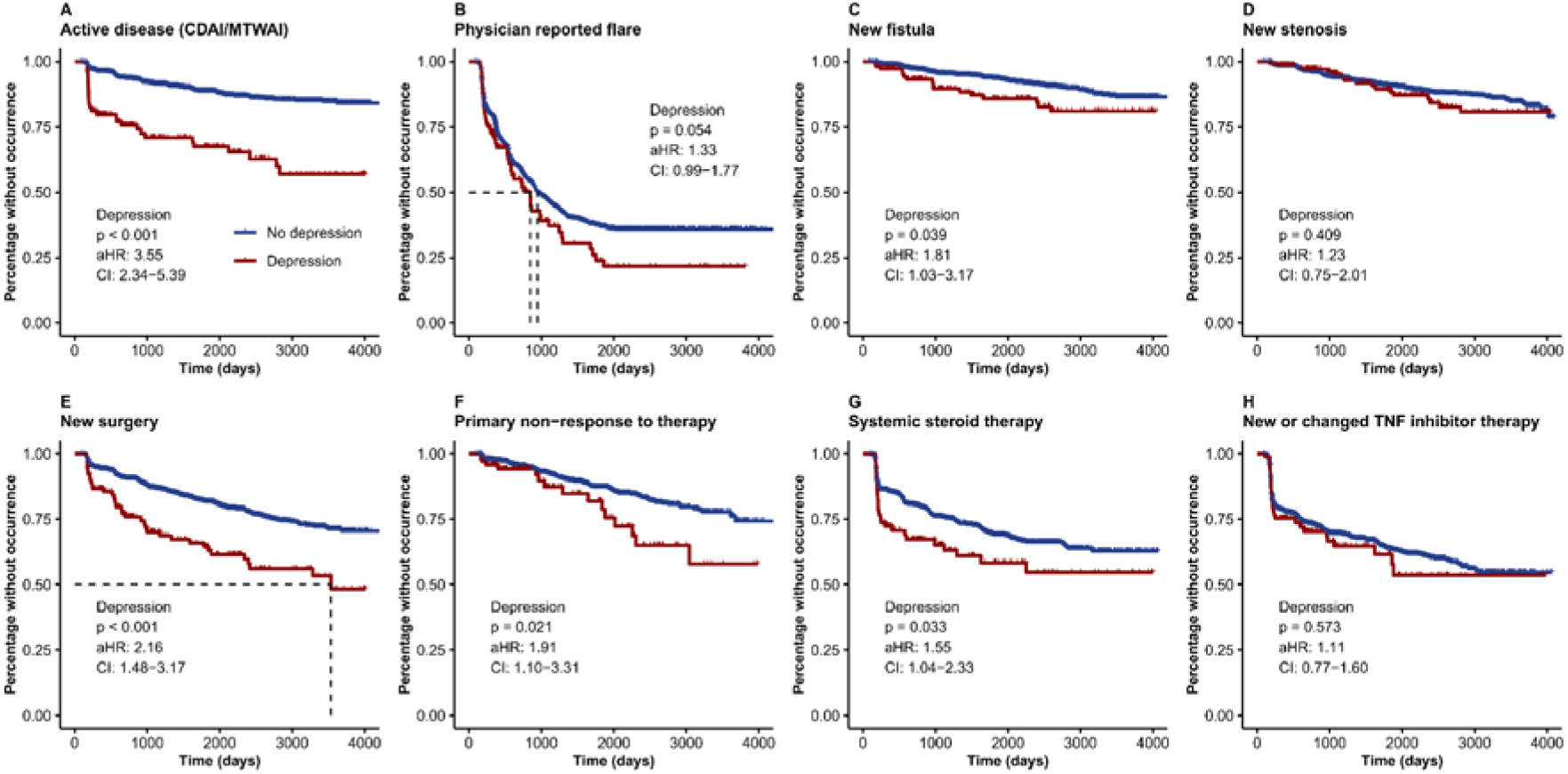
Kaplan Meier curves showing how depression increases the hazards for future adverse outcomes over time. Percentage of IBD patients without experiencing the respective outcome (**A-H**) stratified for depression. Analysis: multivariable Cox proportional hazards models. Abbreviations: HR: hazard ratio, CI: confidence interval, TNF: tumor necrosis factor, CDAI: Crohn’s Disease Activity Index, MTWAI: Modified Truelove and Witts Severity Index.

**Figure 3:**
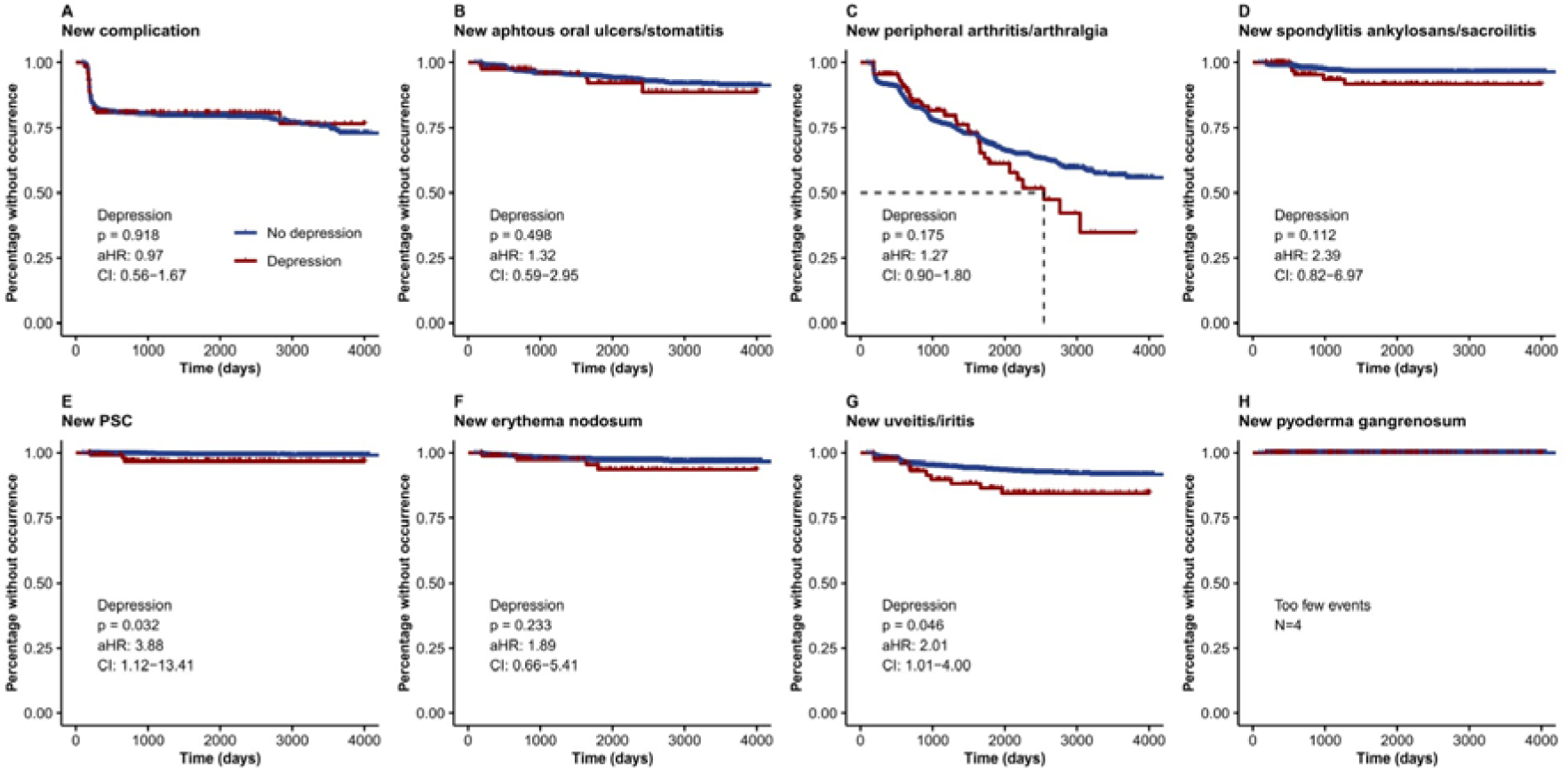
Kaplan Meier curves showing how depression increases the hazards for future EIMs over time. Percentage of IBD patients without experiencing the respective outcome (**A-H**) stratified for depression. Analysis: multivariable Cox proportional hazards models. Abbreviations: HR: hazard ratio, CI: confidence interval, PSC: primary sclerosing cholangitis.

**Figure 4:**
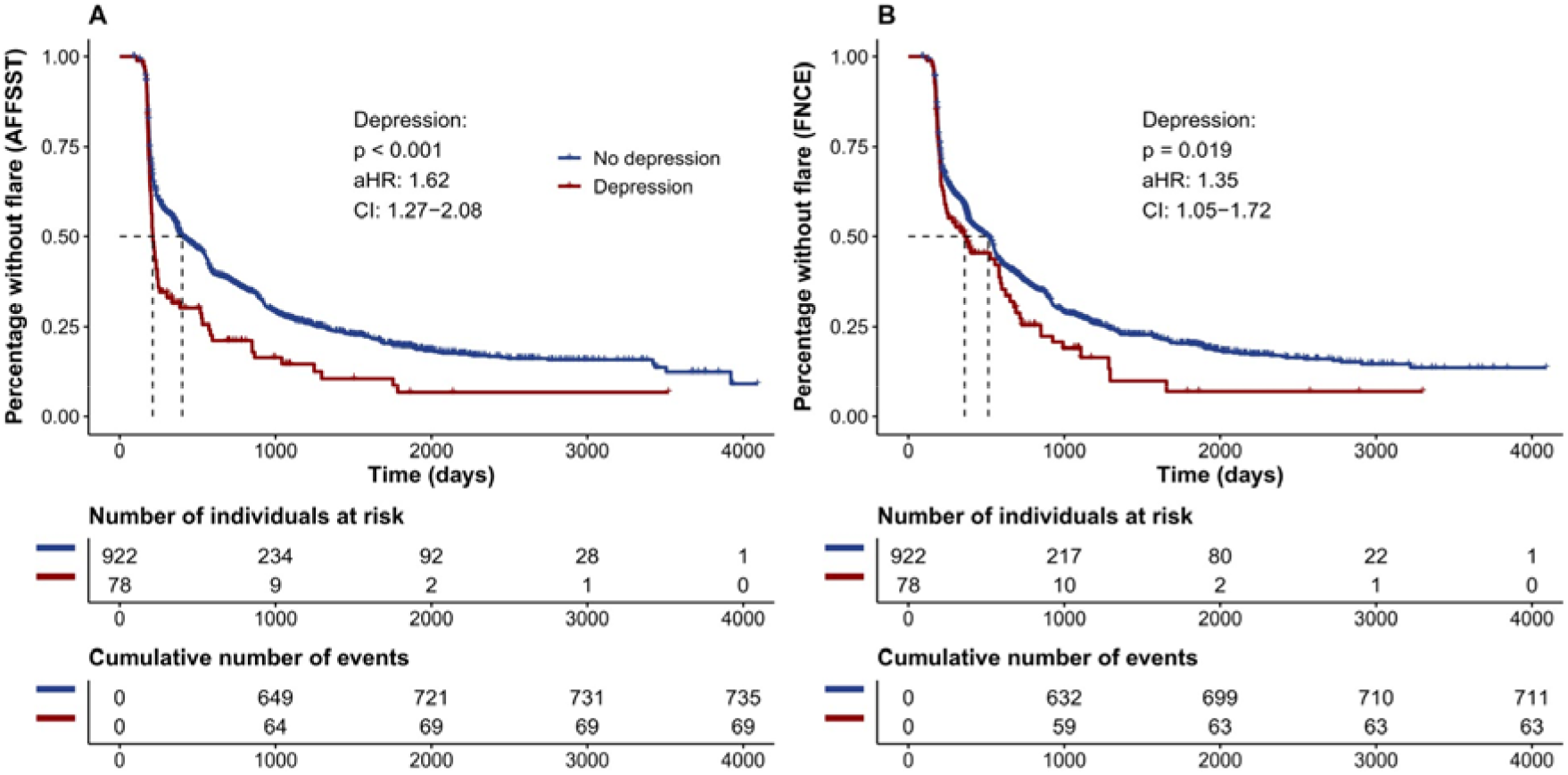
Kaplan Meier curves showing how depression increases the hazard for future clinical deterioration (flare, composite endpoints) over time. **A:** Percentage of IBD patients without a flare according to the AFFSST-definition, stratified according to the presence of depression. **B:** Percentage of IBD patients without a flare according to the FNCE-definition, stratified according to the presence of depression. Analysis: multivariable Cox proportional hazards models. Abbreviations: HR: hazard ratio, CI: confidence interval, IBD: inflammatory bowel disease, AFFSST: active disease, physician reported flare, new fistula, new stenosis, surgery or new systemic therapy, FNCE: physician reported flare, non-response to therapy, new complication or EIM.

### SNPs effects on depression-free survival

To assess the effects of both SNPs on the hazard for continuance or new onset of depressive symptoms (HADS-D ≥11) over time, we implemented Cox proportional hazards models. In this time-to-event analyses, the TC allele combination of rs588765 had significant protective effects regarding depressive symptoms in the univariable model (hazard ratio (HR)): 0.68, confidence interval (CI): 0.46-0.97, p=0.042). The association with rs588765-TC remained robust in the multivariable analysis (adjusted hazard ratio (aHR): 0.67, CI: 0.46-0.97, p=0.035; **Table 4** and **Figure 1; panel A)**. Protective effects of rs588765-TC were independent of disease type and similar for CD and UC patients, although non-significant in each subgroup (CD: aHR: 0.70, p=0.155; UC: aHR: 0.58, p=0.100).

**Table 4.** Cox proportional hazards model - hazard ratios for new or persistent depression among IBD patients over time. **Predictors for new or persistent depression (HADS-D ≥11) over time.** Results are presented as hazard ratios for new or persistent depression among IBD patients over time; p-values are indicated as follows: *: p<0.1, **: p<0.05, ***: p<0.01. 919 patients were analyzed and 114 times the endpoint was reached. Analysis: Cox proportional hazards model. Abbreviations: CI: 95% confidence interval, IBD: inflammatory bowel disease, HADS: Hospital Anxiety and Depression Scale, T: thymine, C: cytosine, UC: ulcerative colitis, IC: indeterminate colitis, CD: Crohn’s disease, BMI: body mass index, CDAI: Crohn’s Disease Activity Index, MTWAI: Modified Truelove and Witts Severity Index, TNF: tumor necrosis factor.

| Endpoint variable: depression (HADS depression sub score $\geq 11$) | | | |
| --- | --- | --- | --- |
|  | Univariable | Multivariable<br>(rs588765) | Multivariable<br>(rs2522833) |
| **rs588765: TC (vs. CC/TT)** | **0.68 (CI: 0.46-0.99)**<br>** | **0.67 (CI: 0.46-0.97)**<br>** |  |
| **rs2522833: number of C alleles (0-2)** | 1.12 (CI: 0.87-1.44) |  | 1.10 (CI: 0.86-1.41) |
| **Sex: male** |  | 0.80 (CI: 0.54-1.19) | 0.79 (CI: 0.54-1.18) |
| **Diagnosis: UC/IC (vs. CD)** |  | 0.82 (CI: 0.51-1.30) | 0.80 (CI: 0.50-1.27) |
| **Time since diagnosis (years)** |  | 0.99 (CI: 0.97-1.01) | 0.99 (CI: 0.97-1.01) |
| **Age (years)** |  | **1.03 (CI: 1.01-1.04)**<br>*** | **1.03 (CI: 1.01-1.04)**<br>*** |
| **BMI** |  | 1.00 (CI: 0.95-1.04) | 1.00 (CI: 0.96-1.04) |
| **Smoking status: smoker** |  | **1.63 (CI: 1.09-2.42)**<br>** | **1.65 (CI: 1.11-2.44)** ** |
| **Daily alcohol consumption: yes** |  | 1.21 (CI: 0.71 -2.07) | 1.24 (CI: 0.73 -2.12) |
| **Disease activity (standardized CDAI/MTWAI)** |  | **1.53 (CI: 1.31-1.79)**<br>*** | **1.52 (CI: 1.30-1.78)**<br>*** |
| **Disease related surgery prior to enrolment** |  | 1.15 (CI: 0.74-1.78) | 1.14 (CI: 0.73-1.78) |
| **Systemic steroids: yes** |  | 0.99 (CI: 0.60-1.63) | 0.99 (CI: 0.60-1.64) |
| **TNF inhibitors: yes** |  | 1.26 (CI: 0.80-1.99) | 1.25 (CI: 0.79-1.96) |
| **Number current therapies** |  | 1.07 (CI: 0.87-1.31) | 1.06 (CI: 0.86-1.30) |

For the C-allele of rs2522833, no significant association was detected (aHR: 1.10, CI: 0.86-1.41, p=0.455, **Table 4** and **Supplementary figure 5; panel A**).

### Depressive symptoms as a hazard for IBD disease course

Patients with depressive symptoms (HADS-D ≥11) had a significantly higher hazard of experiencing active disease (CDAI ≥150/MTWAI ≥10) during follow-up (HR: 3.18, p<0.001, **Table 5** and **Supplementary figure 6; panel A**); this association remained significant after correction for confounders (aHR: 3.25, p<0.001, **Table 5** and **Figure 2; panel A**). We also observed a significant negative impact of depressive symptoms on the endpoints *new surgery* (aHR: 1.92, p>0.001), *systemic steroid therapy* (aHR: 1.55, p=0.033) and *primary non-response to therapy* (aHR: 1.79, p=0.044, **Figure 2; panel E-G**, univariable results: **Supplementary figure 7)**. In other words, patients were more likely to experience the respective endpoints when depressive symptoms were present before, even when corrected for possible confounders.

**Table 5.**
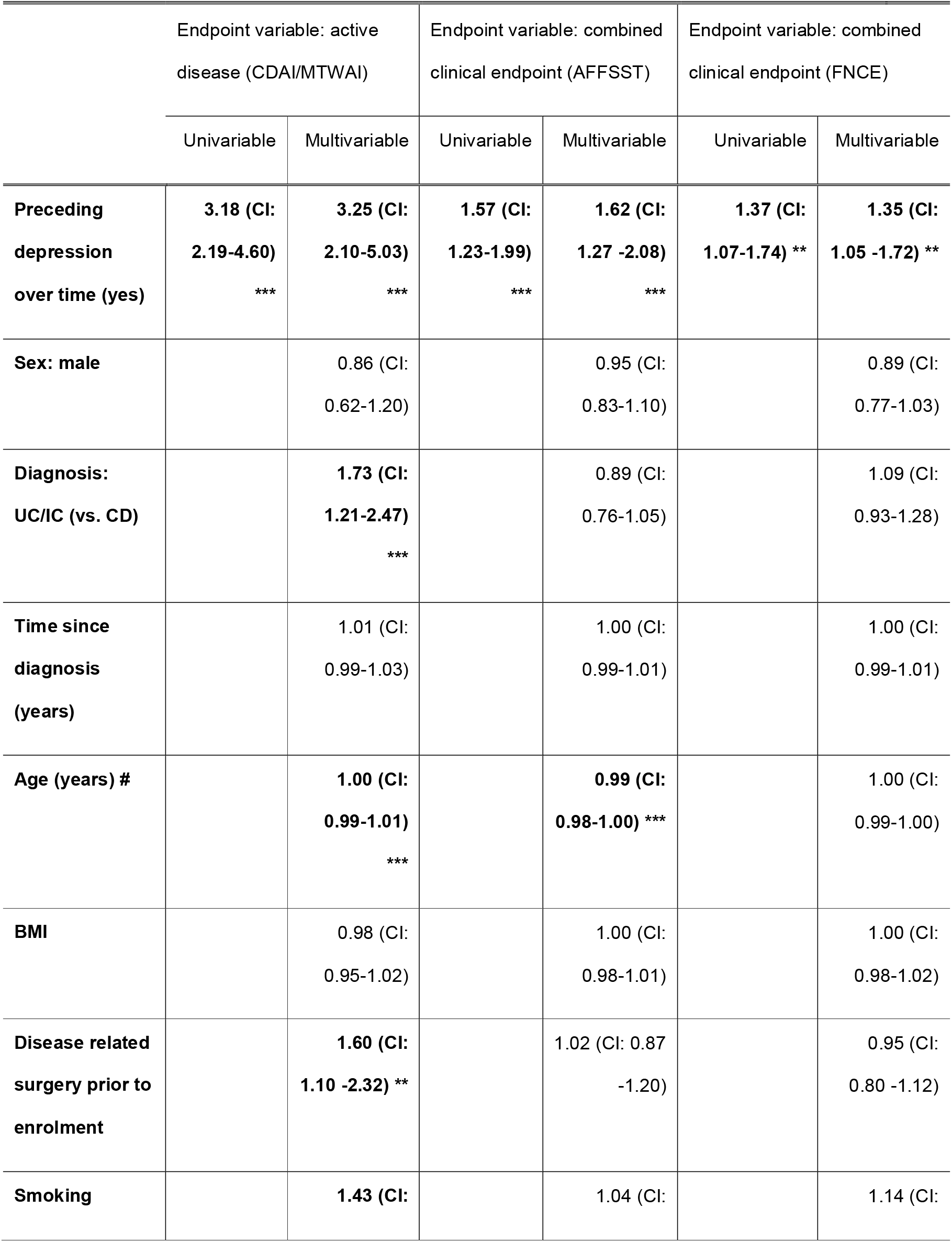
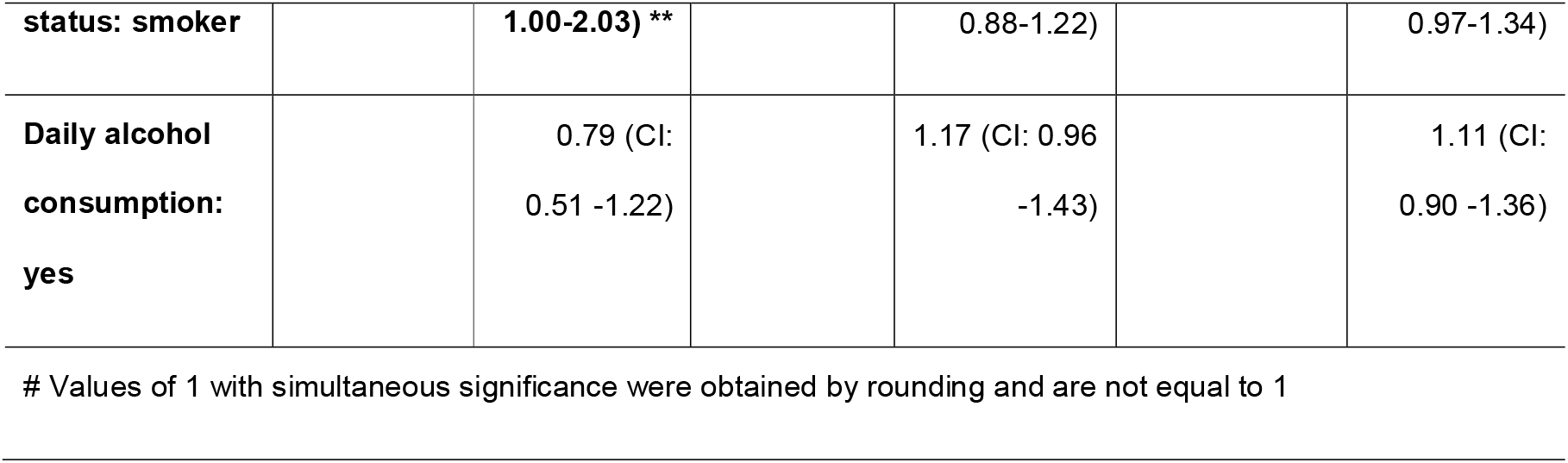
Cox proportional hazards model - hazard ratios for active disease and clinical deterioration (flares) in IBD patients over time. **Predictors for active disease and clinical deterioration (flare) in IBD patients over time.** Results are presented as hazard ratio for active disease or clinical deterioration (flare) respectively, in IBD patients over time. Active disease was defined as CDAI ≥150 or MTWAI ≥10 in CD and UC patients, respectively. A flare was defined by the AFFSST or FNCE composite endpoint; p-values are indicated as follows: *: p<0.1, **: p<0.05, ***: p<0.01. Analysis: Cox proportional hazards model. Abbreviations: CI: 95% confidence interval, IBD: inflammatory bowel disease, HADS: Hospital Anxiety and Depression Scale, UC: ulcerative colitis, IC: indeterminate colitis, CD: Crohn’s disease, BMI: body mass index, CDAI: Crohn’s Disease Activity Index, MTWAI: Modified Truelove and Witts Severity Index, AFFSST: active disease, physician reported flare, new fistula, new stenosis, surgery or new systemic therapy, FNCE: physician reported flare, non-response to therapy, new complication or EIM.

Overall, for all clinical endpoints, patients with depressive symptoms had a higher hazard for clinical deterioration, even though this effect did not reach significance in all cases (**Figure 2**). Furthermore, depressive symptoms also tended to increase the hazard for individual EIMs and significant effects for new PSC (aHR: 3.88, p=0.032) and new uveitis/iritis (aHR: 2.01, p=0.046) in multivariable models were observed (**Figure 3; panel E** and **G**, univariable results: **Supplementary figure 8**).

To further investigate the impact of depressive symptoms on IBD disease course,^10^ we assessed two previously used composite endpoints indicating clinical deterioration: AFFSST^10^ (**a**ctive disease, physician reported **f**lare, new **f**istula, new stenosis, **s**urgery or new **s**ystemic **t**herapy, see methods) and FNCE^21^ (physician reported **f**lare, **n**on-response to therapy, new **c**omplication or **E**IM).

Patients with depressive symptoms were also more likely to reach these composite endpoints: in the univariable analysis, the AFFSST endpoint (HR: 1.57, CI: 1.23-1.99, p<0.001, **Table 5**) as well as the FNCE endpoint (HR: 1.37, CI: 1.07-1.74, p=0.012, **Table 5**) were significantly associated with preceding depressive symptoms. Correction for control variables barely changed results and significance was maintained (**Figure 4**).

### Effects of SNPs on IBD activity over time

We tested whether rs588765-TC or rs2522833’s C allele could be directly implicated as protective or risk factors for clinical deterioration (**Table 6**). In univariable models, the TC allele combination of rs588765 showed significant protective effects against active disease (CDAI ≥150/ MTWAI ≥10, HR: 0.78, CI: 0.61-0.99, p=0.042). These significant protective effects of rs588765’s TC alleles remained significant after controlling for confounders (**Figure 1; panel B**).

**Table 6.**
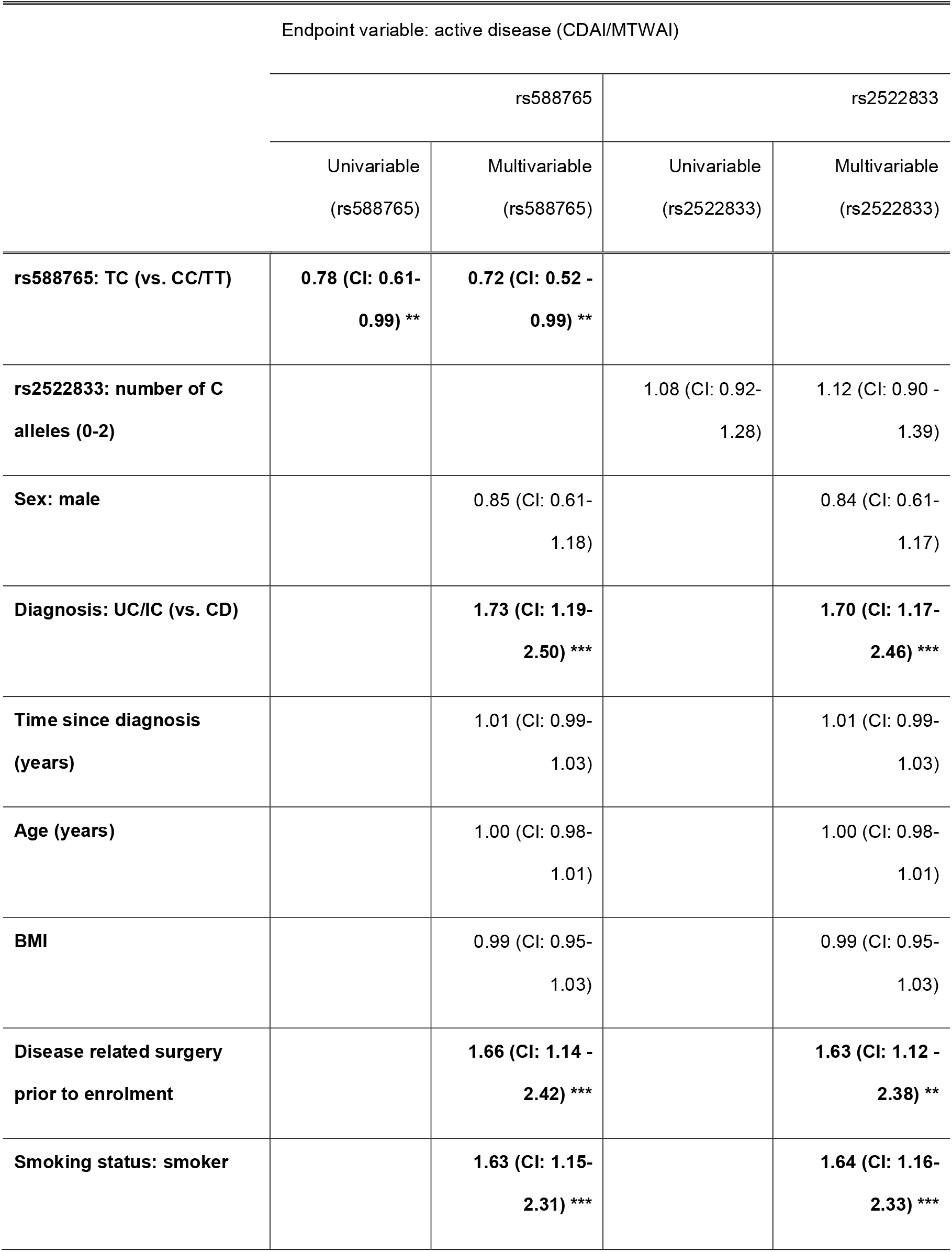
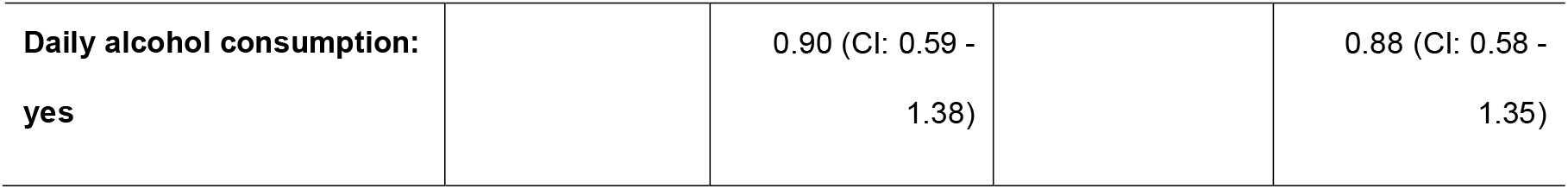
Cox proportional hazards model - hazard ratios for active disease in IBD patients over time. **Predictors for active disease in IBD patients over time.** Results are presented as hazard ratio for active disease in IBD patients over time. Active disease was defined as CDAI ≥150 or MTWAI ≥10 in CD and UC patients, respectively; p-values are indicated as follows: *: p<0.1, **: p<0.05, ***: p<0.01. Analysis: Cox proportional hazards models. Abbreviations: CI: 95% confidence interval, IBD: inflammatory bowel disease, CDAI: Crohn’s Disease Activity Index, MTWAI: Modified Truelove and Witts Severity Index, C: cytosine, T: thymine, UC: ulcerative colitis, IC: indeterminate colitis, CD: Crohn’s disease, BMI: body mass index.

In contrast, the C allele of rs2522833 was not a significant risk factor for active disease (univariable: HR: 1.08, CI: 0.92-1.28, p=0.344, multivariable: **Supplementary figure 5; panel B**).

### Genetic inheritance patterns of rs588765 and rs2522833

In a sensitivity analysis we examined the impact of different allele variants of rs588765 on outcomes of interest. Similarly as for initial screening, the overdominance model (TC vs. TT or CC) seemed to provide the best fit for Cox proportional models with depressive symptoms over time and active disease as endpoints (**Supplementary tables 3** and **4, Supplementary figure 8; panel A** and **C**). Similarly, for rs2522833, the log-additive inheritance model which uses the number of C alleles (0, 1 or 2) seemed to be the best fit regarding active disease (**Supplementary table 4, Supplementary figure 8; panel D**), but not regarding depressive symptoms over time (**Supplementary table 3, Supplementary figure 8; panel B**). No formal statistical testing comparing the fits of genetic models was done.

## Discussion

In our cross-sectional analysis of depressive symptoms in IBD patients, we identified variants of two SNPs (rs588765-TC, rs2522833’s C allele) that were significantly (q<0.10) associated with depression in IBD. The presence of depressive symptoms was a strong risk factor for subsequent clinical deterioration in time-to-event analyses, confirming and extending earlier findings by showing robust negative effects on numerous clinical endpoints as well as two previously published composite endpoints for clinical deterioration.^10,21^ Finally, we could demonstrate that the TC allele of rs588765 was protective regarding depression and IBD activity in a time-to-event analysis.

IBD patients are a high-risk group for developing depression.^6^ In line with these findings, we observed an elevated point prevalence of depressive symptoms in our cohort (77/814, 9.5% among CD patients and 40/618, 6.5% among UC/IC patients) compared to the general population. For instance, point prevalence in a non-clinical Swiss sample with comparable assessment was estimated to be only 3.1%,^40^ similar to findings in the USA^41^ and the UK.^26^

We found a significant negative association between depressive symptoms and patients with a specific variant in one of the preselected SNPs (rs588765-TC, ORs of 0.43, q=0.05) in a cross-sectional analysis with enrolment data. This is an observation that, to the best of our knowledge, had not been described before. Moreover, the time-to-event analysis indicated a protective effect of the same allele combination regarding new or persistent depressive symptoms over time (aHR: 0.67, p=0.035), thus independently confirming protective properties of rs588765-TC. Finally, protective effects of rs588765-TC regarding depression translated to protection regarding IBD flares (CDAI ≥150/MTWAI ≥10, aHR: 0.72, p=0.045). All analyses were corrected for smoking status and alcohol consumption indicating that protection regarding depression and flares is independent of these features.

Rs588765 is part of the chr15q25.1 region and represents a well-established tag-SNP on the CHRNA5-CHRNA3-CHRNB4 gene cluster^42,43^ that encodes the nAChR subunits α5, α3, and β4. Rs588765 is located in the neuronal acetylcholine receptor subunit α5 gene (CHRNA5).^44^ CHRNA5’s gene product serves as a subunit of pentameric nAChRs.^45^ Rs588765 impacts the expression of CHRNA5 in human brain tissue^46^ with subjects homozygous with two minor alleles (TT) showing the highest CHRNA5 mRNA expression.^46^ NAChRs are ligand-gated ion channels that mediate signal transmission at synapses and modulate the release of different neurotransmitters.^47^ Addition of an α5 subunit to nAChRs α4β2, the most frequent receptor subtype in the brain, increases the rate of receptor desensitization and calcium permeability.^47^ Similarly to the brain, CHRNA5 seems to mediate excitatory nicotinic cholinergic transmission in the enteric nervous system (ENS).^48^ Rs588765 had been associated with nicotine and alcohol dependence,^15,16^ also among SIBDC patients.^17^ Moreover, the associated CHRNA5-CHRNA3-CHRNB4 region has also been suggested to contain risk genes for affective disorders such as major depressive disorder ^49-51^ and recent findings suggest that nAChRs in general are involved in psychiatric disorders such as depression.^14^

The genetic model of overdominance we stated best fitting for rs588765 refers to advantageous effects of the heterozygous variant compared to homozygous variants.^52^ Examples include protective effects of hemoglobin mutation against malaria of in human sickle cell anaemia.^53^ For rs588765, functional interactions of genetic variants associated with the T and C allele, respectively, seem possible as a variant of epistasis. Another plausible explanation for overdominance is the concept of an inverted U-shaped response curve.^54^

The second SNP (rs2522833) associated with depressive symptoms is part of the piccolo presynaptic cytomatrix protein (PCLO) gene. The presynaptic cytomatrix describes a complex electron-dense meshwork of proteins involved in transmitter release.^55,56^ Moreover, PCLO has been implicated to potentially contributing to short-term neuronal plasticity.^48^ The PCLO gene has been associated with depressive conditions such as major depressive disorder^57^ and the C allele of rs2522833 specifically might increase an individual’s vulnerability regarding depression.^58^ Our current results align well with these earlier findings and the C allele was a significant (q<0.1) log-additive risk factor for depressive symptoms at baseline (ORs: 1.73, q=0.059). However, our study did not detect significance for rs2522833 in the time-to-event analyses.

In our analysis, depressive symptoms remained a strong and highly significant risk factor for future clinical deterioration according to several clinical measures including the well-established disease scores CDAI and MTWAI and two pre-defined, partially overlapping composite flare definitions (AFFSST^10^ and FNCE^21^, respectively). Depressive symptoms also increased the likelihood for future occurrence of nearly all other IBD-related endpoints tested over time (Figures 2 and 3), even though without uniform statistical significance. This suggests that the specific definition for a flare is largely irrelevant for the association with depression.

A growing body of evidence indicates that stress in general^11,21^ and depressive symptoms specifically^10^ are associated with clinical flares in IBD patients. Whether the psychological state of an IBD patient can indeed directly increase gut inflammation remains an important question in IBD research.^59^ Our results support a direct impact of depressive symptoms on gut inflammation: i) we show strong associations between rs588765, depressive symptoms and IBD flares (Tables 3-6); ii) depressive symptoms remain a strong risk factor for flares according to various definitions (CDAI/MTWAI: aHR: 3.25, p<0.001, AFFSST: aHR: 1.62, p<0.001, FNCE: aHR: 1.35, p=0.019) and iii) the TC allele of rs588765 with protective effects regarding depressive symptoms (aHR: 0.67, p=0.035) was also protective against IBD flares (CDAI/MTWAI: aHR: 0.72, p=0.045). The combination of clinical and genetic data strengthens this point; however, indirect effects cannot be entirely excluded. For instance, depression might be a marker for subclinical inflammation or might affect reporting of symptoms by patients. Another interesting possibility could be an effect of rs588765-TC on CHRNA5 expression in the gut and the brain simultaneously, thus independently explaining depressive symptoms and flares. Further mechanistic studies would be necessary to better understand our observation.

Our study has several strengths and limitations. Strengths include the excellent clinical characterization regarding IBD-related outcomes and the sequential measurements of the key variable *depressive symptoms* and the clinical endpoint variables over a long time enabling the combination of cross-sectional and time-to-event analyses. Limitations are the relatively small sample size for genetic analyses, yielding in lower statistical power for some analyses. Moreover, missing data points, e.g. for therapy or surgery data, further reduced statistical power for some analyses. Furthermore, even though serial measurements for HADS-D were available and a HADS-D score of ≥11 represents a well-established cut off strongly indicating depression,^23-26^ a diagnosis of depression normally requires direct clinical assessment by a psychiatrist. Furthermore, for validation of our data, independent confirmation in another IBD cohort would have been desirable. Finally, in 2019, a large meta-analysis for the risk of depression including 807,553 individuals was published. This study identified 102 independent variants (not rs588765, rs2522833) of which 87 were replicated in an independent sample.^13^ Therefore, future studies with a larger number of SNPs will now be possible.

In conclusion, our study confirms an intimate relationship between depressive symptoms and IBD disease course. The prevalence of depression among IBD patients is high and active disease further increases the hazard for depression.^6^ Vice versa, depressive symptoms also independently increased the hazard for IBD flares over time. Moreover, two SNPs (rs588765-TC, rs2522833’s C allele) were associated with depressive symptoms among IBD patients and one of those SNPs (rs588765-TC) was also associated with high IBD activity. These findings are in accordance with a multidirectional relationship between genetics, depression and IBD disease course. Therefore, a comprehensive therapeutic concept for IBD patients, targeting inflammation and depression might further improve patient care.^60^

## Data Availability

All data is available and in possession of the corresponding author.

## Acknowledgments

The authors would like to thank SIBDC patients and SIBDC staff for their commitment and are grateful to the SIBDCS Scientific Committee for granting approval for this study.

## Abbreviations

aHR: adjusted hazard ratio
AIC: Akaike information criterion
BMI: body mass index
CD: Crohn’s disease
CDAI: Crohn’s Disease Activity Index
CHRNA5: cholinergic receptor nicotinic alpha 5 subunit
CI: confidence interval
EIM: extraintestinal manifestations
ENS: enteric nervous system
HADS: Hospital Anxiety and Depression Scale
HR: hazard ratio
IBD: inflammatory bowel disease
IC: indeterminate colitis
LD: linkage disequilibrium
MTWAI: Modified Truelove and Witts Severity Index
NAChR: nicotinic acetylcholine receptors
OR: odds ratio
PCLO: piccolo presynaptic cytomatrix protein
PSC: primary sclerosing cholangitis
R^2^: square of the correlation coefficient
SIBDC: Swiss IBD cohort
SNP: single nucleotide polymorphism
t_n_: follow-up time *n* of recording
TNF: tumor necrosis factor
UC: ulcerative colitis

